# Glaucoma home-monitoring using a tablet-based visual field test (Eyecatcher): An assessment of accuracy and adherence over six months

**DOI:** 10.1101/2020.05.28.20115725

**Authors:** Pete R. Jones, Peter Campbell, Tamsin Callaghan, Lee Jones, Daniel S. Asfaw, David F. Edgar, David P. Crabb

## Abstract

**Purpose:** To assess accuracy and adherence of visual field (VF) home-monitoring in a pilot sample of glaucoma patients.

**Design:** Prospective longitudinal observation.

**Methods:** Twenty adults (median 71 years) with an established diagnosis of glaucoma were issued a tablet-perimeter (Eyecatcher), and were asked to perform one VF home-assessment per eye, per month, for 6 months (12 tests total). Before and after home-monitoring, two VF assessments were performed in-clinic using Standard Automated Perimetry (SAP; 4 tests total, per eye).

**Results:** All 20 participants could perform monthly home-monitoring, though one participant stopped after 4 months (Adherence: 98%). There was good concordance between VFs measured at home and in the clinic (*r* = 0.94, *P* < 0.001). In 21 of 236 tests (9%) Mean Deviation deviated by more than ±3*dB* from the median. Many of these anomalous tests could be identified by applying machine learning techniques to recordings from the tablets’ front-facing camera (Area Under the ROC Curve = 0.78). Adding home-monitoring data to 2 SAP tests made 6 months apart reduced measurement error (between-test measurement variability) in 97% of eyes, with mean absolute error more than halving in 90% of eyes. Median test duration was 4.5*mins* (*Quartiles*: 3.9−5.2*mins*). Substantial variations in ambient illumination had no observable effect on VF measurements (*r* = 0.07, *P* = 0.320).

**Conclusions:** Home-monitoring of VFs is viable for some patients, and may provide clinically useful data.

## 1. Introduction

People with glaucoma, or at risk of developing glaucoma, require lifelong monitoring, including periodic (e.g., annual^1^) visual field (VF) examinations^2^. The volume of outpatient appointments required (> 1 million/year in the UK alone^3^) is placing glaucoma services under increasing strain: as evidenced by a growing appointment backlog^4^ and cases of avoidable sight loss due to treatment delays^5,6^. Globally, the challenge of glaucoma management is only likely to intensify over the coming decades^7^, with aging societies^8,9^ and calls for increased monitoring^1^ and earlier detection^10^. Hospital assessments also cannot be performed with the frequency required for best patient care. Multiple studies have shown that intensive VF monitoring could help to identify and prioritize individuals most at risk of debilitating sight loss^11–15^ (i.e., younger patients with fast-progressing VF loss^16^). And frequent (e.g., monthly) monitoring is likely to be of particular benefit for those patients for whom rapid progression is most likely (e.g., optic disk hemorrhage patients^17–19^) or most costly (e.g., monocular patients^20^).

In short, the *status quo* of hospital-only VF monitoring is costly and insufficient. The solution may lie in home-monitoring^14,21,22^. By collecting additional VF data between appointments, hospital visits could be shortened, and in low-risk patients reduced in frequency or conducted remotely: decreasing demand on outpatient clinics. Home-monitoring would further allow for more VF testing, and more frequent VF testing: both important for rapid, robust clinical decision-making^12,23^. For these reasons, interest in home monitoring is growing both for glaucoma^14,21,22^ and the management of other chronic ophthalmic conditions^24–27^, as well as in healthcare generally^28^. This interest is likely to intensify following COVID-19, as hospitals look to minimize outpatient appointments^29^.

Technological advances mean VF home-monitoring is now a realistic proposition. Several portable perimeters have been developed that use ordinary tablet-computers (e.g., Melbourne Rapid Fields^30–32^; Eyecatcher^33^) or head-mounted displays (e.g., imo^34,35^, Mobile Virtual Perimetry^36^). Such devices are small and inexpensive enough for patients to take home, and several appear capable of approximating conventional SAP when operated under supervision^31,37,38^.

What remains unclear is whether VF home-monitoring works in practice. Are glaucoma patients willing and able to comply with a home-testing regimen (adherence), and do ‘personal perimeters’ continue to produce high-quality VF data when operated at home, unsupervised (accuracy)?

To investigate these questions, twenty people with established glaucoma were given a table perimeter (Eyecatcher) to take home for 6 months. They were asked to perform one VF assessment a month in each eye. Accuracy was assessed by comparing measurements made at home to conventional SAP assessments made at the study’s start and end. Adherence was quantified as the percentage of tests completed. Eyecatcher is not yet available for general use, however the source-code is freely available online.

To reflect the likely clinical reality of home-monitoring, we used inexpensive and commonly available hardware (∼£350 per person). Ten participants were given no practice with the test before taking it home. The other 10 performed the test once in each eye under supervision. During home-testing, the tablet-computer’s forward-facing camera recorded the participant. This allowed us to confirm the correct eye was tested, to record variations in ambient illumination, and to investigate whether affective computing techniques (e.g., head-pose tracking and facial-expression analysis) could identify suspect tests^39^.

## 2. Methods

### 2.1. Participants

Participants were 20 adults (10 female) aged 62 − 78 years (Median: 71), with an established diagnosis of: primary open angle glaucoma (*N* = 18, incl. 6 normal tension), angle closure glaucoma (*N* = 1), or secondary glaucoma (*N* = 1). Participants lived across south England and Wales (see **Supplemental Figure S1**), and were under ongoing care from (different) consultant ophthalmologists. Participants were the first 20 respondents to an advertisement placed in the International Glaucoma Association newsletter (*IGA News*: https://www.glaucoma-association.com/ about-the-iga/what-we-do/magazine), and were assessed by a glaucoma-accredited optometrist (author PC) who recorded: ocular and medical histories, logMAR acuity, and standard automated perimetry using a Humphrey Field Analyzer 3 (HFA; Carl Zeiss Meditec, CA, US; Swedish Interactive Threshold Algorithm (SITA) Fast; 24–2 grid). All patients exhibited best corrected logMAR acuity < 0.5 in the better eye, and none had undergone ocular surgery or laser treatment within six months prior to participation. Severity of visual field loss in the worse eye, as measured by HFA Mean Deviation (MD), varied from −2.5 *dB* (early loss^40^) to −29.9 *dB* (advanced), although the majority of eyes exhibited moderate loss (Median: −8.9 *dB*). All HFA assessments (4 per eye) are shown in the *Results*, and all exhibited a False Positive rate below 15% (Median: 0%).

Written informed consent was obtained prior to testing. Participants were not paid, but were offered travel expenses. The study was approved by the Ethics Committee for the School of Health Sciences, City, University of London (#ETH1819–0532), and carried out in accordance with the tenets of the Declaration of Helsinki.

### 2.2. Procedure

As shown in **Figure 1A**, participants were asked to perform one VF home-assessment per eye, per month, for 6 months (12 tests total). Beforehand, participants attended City, University of London, where they were issued with the necessary equipment, including: a tablet computer (**Fig 1B**), an eye-patch, screen wipes, and a brief set of written instructions. Ten participants (50%) were also randomly selected to practice the Eyecatcher test once in each eye, under supervision. All participants performed two HFA assessments in each eye (24–2 SITA Fast).

**Fig 1.**
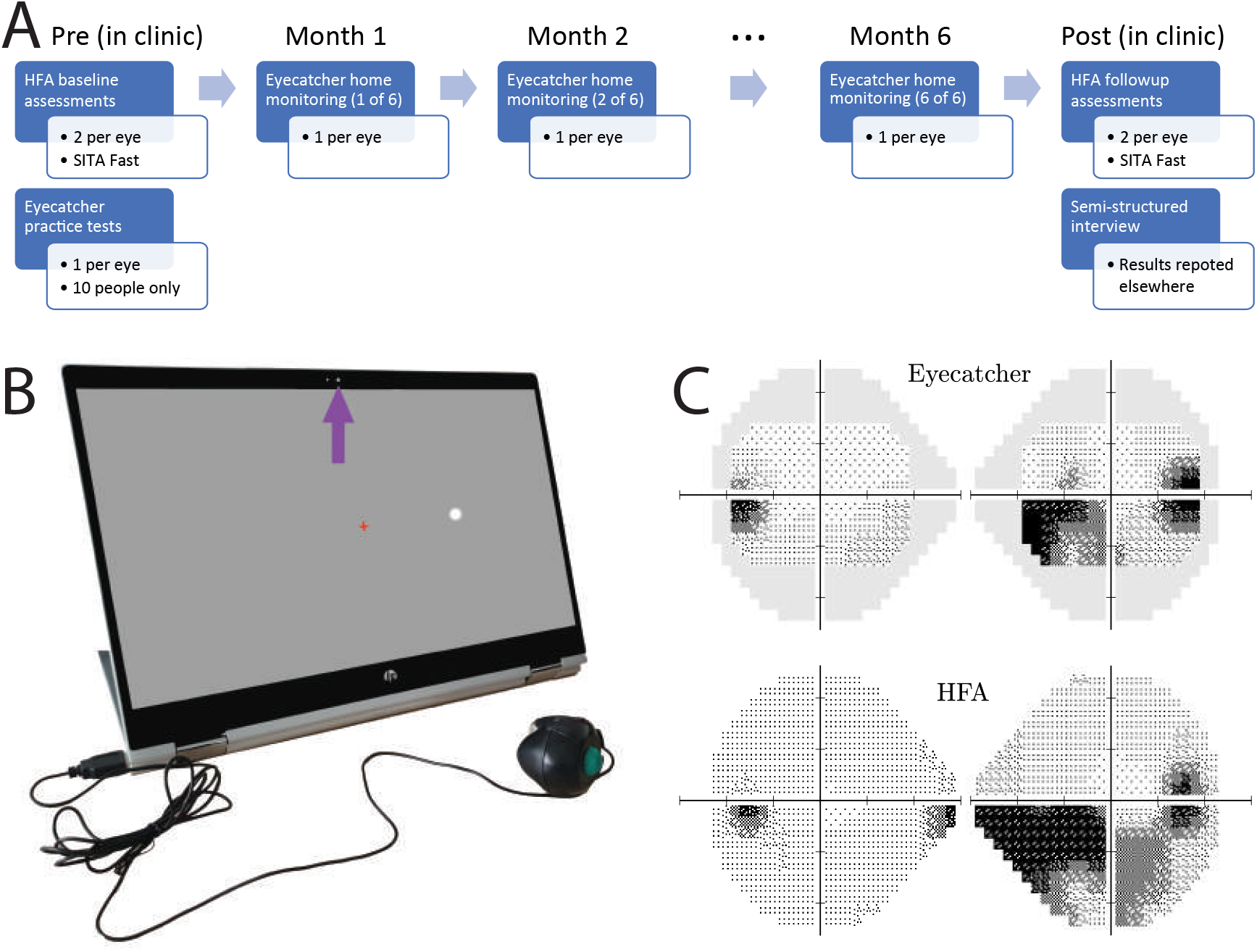
Methods. **(*A*)** Study timeline. **(*B*)** Hardware: Home perimetry was performed using an inexpensive tablet-perimeter (‘Eyecatcher’). During each Eyecatcher assessment, live recordings of the participant were made via the screen’s front facing camera (purple arrow). Participants were asked to fixate the central red cross throughout, and to press the button when a white dot was seen **(*C*)** Output: Example measures of visual field loss from a single participant, with same-patient data from the HFA for comparison. Greyscales were generated using MATLAB code available at: https://github.com/petejonze/VfPlot.

During the 6-month home-testing period, participants had access to support via telephone and email, and received an email reminder once a month when the test was due. After the home-monitoring period was complete, participants returned to City, University of London, and again performed two HFA assessments in each eye. They also completed a semi-structured interview, designed to assess the acceptability of home monitoring, and to identify any potential barriers to use. A qualitative assessment of these interviews will be reported elsewhere. One participant (ID16) was unable to return due to the COVID-19 quarantine. They instead mailed their computer, and performed their interview via telephone. No follow-up HFA assessment could be performed in this individual, but given their ocular history their VF is expected to have been stable.

### 2.3. The Eyecatcher visual field test

Visual fields were assessed using a custom screen-perimeter (**Fig 1B**), implemented on an inexpensive HP Pavilion x360 39.6 cm (15.6”) tablet-laptop (HP Inc., Paolo Alto, CA, United States). The test was a variant of the ‘Eyecatcher’ visual field test: described previously^33,38^ and freely available online (https://github.com/petejonze/Eyecatcher). It was modified in the present work to more closely mimic conventional static threshold perimetry; most notably by: employing a ZEST thresholding algorithm^41^, a central fixation-cross, and a button press response. The software was implemented in MATLAB using Psychtoolbox v3^42^, and used bit-stealing to ensure > 10-bit luminance precision^43^. Extensive photometric calibration was performed on each device to ensure stimulus uniformity across the display (see [^44^] for technical details).

During the test, participants were asked to fixate a central cross, and to press a button when they saw a flash of light (Goldmann III dots with Gaussian-ramped edges). Unlike conventional perimetry, participants received visual feedback (a ‘popping’ dot) at the true stimulus location after each button press. This feedback was intended to keep participants motivated and alert during testing and was generally well-received by participants, though 4 reported being sometimes surprised when feedback appeared at an unexpected location.

Testing was performed monocularly (fellow eye patched). The right eye was always tested first, and participants could take breaks between tests. Participants were asked to position themselves 55 cm from the screen (a distance marked on the response-button cable), and to perform the test in a dark, quiet room. In practice, we had no control over fixation stability, viewing distance, or ambient lighting. In anticipation that these may be important confounding factors, participants were recorded during testing using the tablet’s front facing camera (see *Results*).

As shown in **Fig 1C**, the output of each Eyecatcher assessment was 4 × 6 grid of differential light sensitivity (DLS) estimates, corresponding to the central 24 locations of a standard 24–2 perimetric grid (*±*15° horizontal; *±*9° vertical). For analysis and reporting purposes, these values were transformed to be on the same decibel scale as the HFA 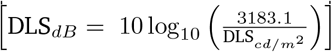 Due to the limited maximum-reliable luminance of the screen (175*cd*/*m*^2^), the measurable range of values was 12.6 *dB* to 48 *dB* (HFA *dB* scale). Sensitivities below 12.6 *dB* could not be measured, and were recorded as 12.6 *dB*. Note, it has been suggested that with conventional SAP, measurements below ∼ 15 *dB* are unreliable and of limited utility^45–47^.

The MRF iPad app has shown promising results under laboratory settings^31^, and was considered for the present study. We chose to use our opensource Eyecatcher software primarily for practical reasons (i.e., we were familiar with it, and could modify it to allow camera recordings and individual screen calibrations).

### 2.4. Analysis

Where appropriate, and as indicated in the text, pointwise DLS values from the HFA were adjusted for parity with Eyecatcher by setting estimated sensitives below 12.6 *dB* to 12.6 *dB*. MD values were then recomputed using age-corrected normative values^48^, using only the central 22 locations tested by both devices (ignoring the two blind spots). Non-adjusted MD values, as reported by the HFA device itself, are also reported in the *Results*.

## 3. Results

**Figure 2** shows Mean Deviations (MD) for all eyes/tests. Adherence (percentage of tests completed) was 98.3%. Nineteen of 20 individuals completed the full regimen of 6 home-monitoring sessions. Participant 20 discontinued home-testing after 4 sessions/months following consultation with the study investigators. This was due to the test exacerbating chronic symptoms of vertigo (also experienced following SAP).

**Fig 2.**
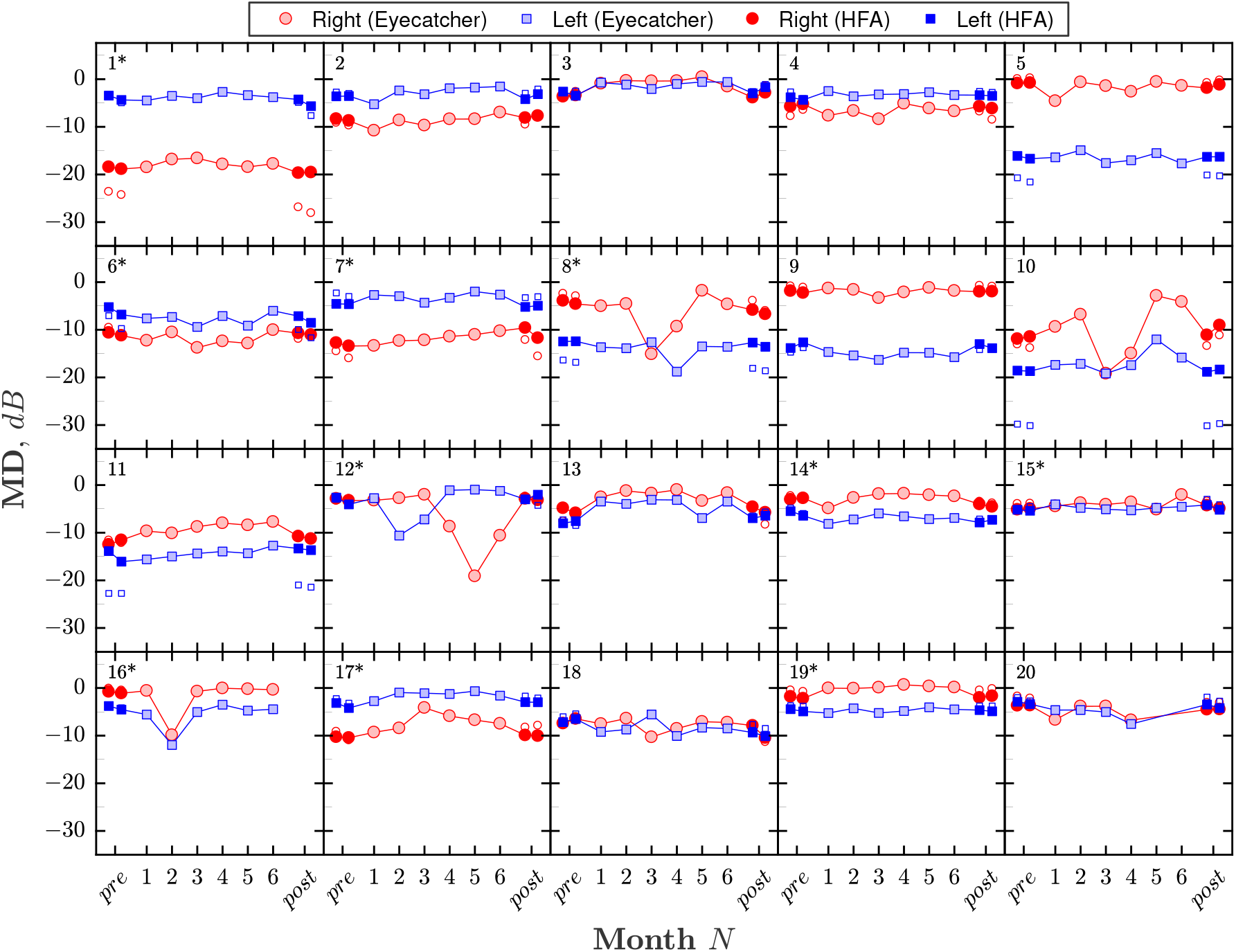
Summary of visual field loss (Mean Deviation; MD) for all eyes/tests. Each panel shows a single participant. Numbers in the top-left of each panel give participant ID, with asterisks denoting the 10 individuals who received initial practice with Eyecatcher. The right eye (*red circles*) was always tested first, followed by the left eye (*blue squares*). Light-filled markers show the results for monthly Eyecatcher home-monitoring assessments. Dark-filled markers show the results of two HFA pre-tests and two HFA post-tests (all tests performed consecutively, same day). For parity, HFA values were computed using only the same 22 (paracentral) test locations as Eyecatcher, and any estimated sensitivities below 12.6 *dB* were set to 12.6 *dB* (to reflect the smaller dynamic range of the Eyecatcher test). Small unfilled markers show the unadjusted MD values as reported by the HFA (i.e., using all 52 test points, and using the full dynamic range). These unfilled markers are most visible (i.e., deviated from the adjusted values) only when field loss was severe. Note that participant 20 chose not complete the final two home monitoring tests, while participant 16 was unable to perform the final HFA assessments due to COVID-19 (see *Body Text* for details).

MD scores were strongly associated between VFs measured at home (mean of 6 Eyecatcher tests) and those measured in the lab (mean of 4 HFA tests), with a correlation of *r*_38_ = 0.94 [**Fig 3A**; *Pearson Correlation*; *P* < 0.001] and a 95% Coefficient of Repeatability of ±3.4 *dB* (**Fig 3B**). For reference, mean agreement between random pairs of HFA assessments was 2.2 *dB* (*CI*_95_: 1.8 − 2.6 *dB*; 20,000 random samples). As shown in **Figure 4**, there was also good concordance between individual VF locations [*Pearson Correlation*; *r*_878_ = 0.86, *P* < 0.001].

**Fig 3.**
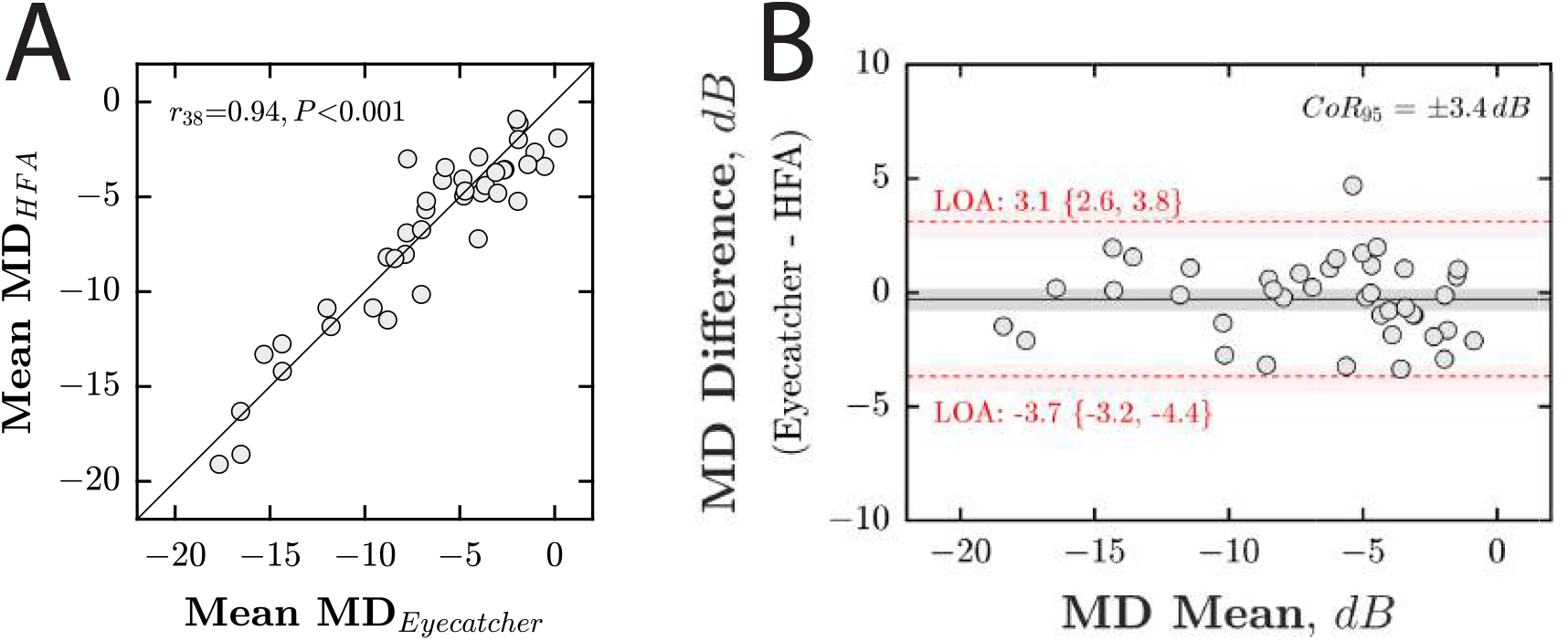
Accuracy (concordance with HFA). **(*A*)** Scatter plot, showing mean MD from the HFA (averaged across all 4 tests), against mean MD from Eyecatcher (averaged across all 6 home tests). Each marker represents a single eye. The solid diagonal line indicates unity (perfect correlation). Statistics show the results of a Pearson correlation. Note that the HFA MD values shown here were adjusted for parity with Eyecatcher’s measurable range/locations (see *Methods*). If the unadjusted raw MD values were used, the correlation was *r*_38_ = 0.91, *P* < 0.001. **(*B*)** Bland-Altman agreement. Red horizontal dashed lines denote 95% Limits of Agreement, with 95% confidence intervals derived using Bootstrapping (Bias-corrected accelerated method, *N* = 20, 000). The 95% Coefficient of Repeatability (CoR_95_) was 3.4 *dB*.

**Fig 4.**
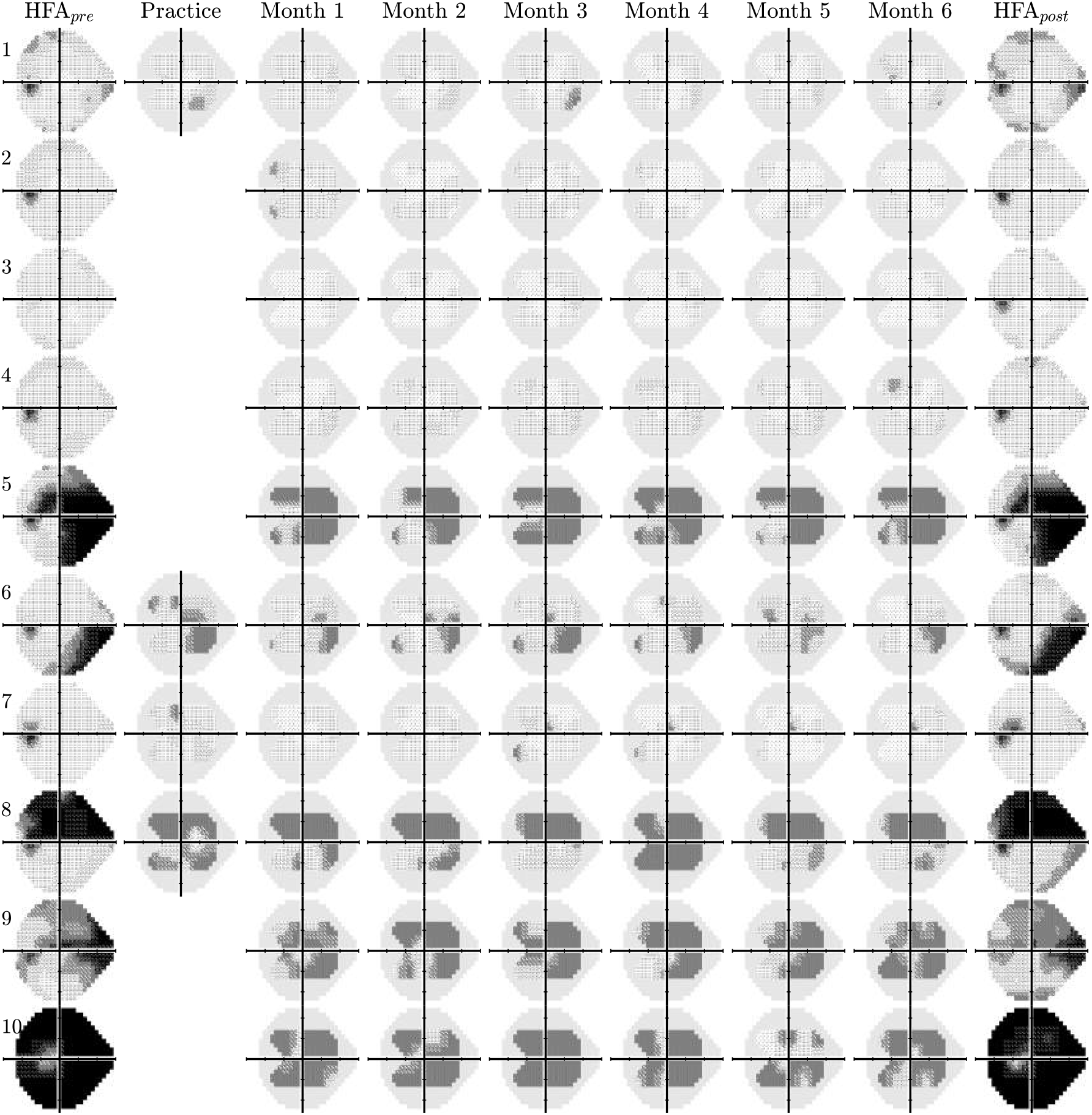
Raw visual field results for 10 randomly selected left eyes (see **Supplemental Figures S2–4** for the other 30 eyes). The first and last columns show mean-averaged data from two ‘pre’ and two ‘post’ reference tests, performed in clinic using a Humphrey Field Analyzer 3 (24–2, SITA Fast). The solid grey regions in the Eyecatcher plots denote those regions of the 24–2 grid not tested due to limited screen size. Only half of participants were randomly selected to complete a supervised practice test (*column 2*).

Some individual tests produced implausible data (e.g., **Fig 2**: *ID8 test 3, ID12 test 5*). In total, there were 21 tests (9%) where MD deviated by more than ±3 *dB* from the average (median of all 6 tests). Of these, 13 (62%) occurred in the right eye (tested first), and 7 (33%) deviated by more than ±6 *dB*. As described in **Supplemental Figure S5**, these statistical outliers could be identified with reasonable sensitivity/specificity [*Area Under the ROC Curve*: 0.78] by applying machine learning techniques to recordings from the tablets’ front-facing camera.

To quantify the extent to which regular home monitoring reduced VF measurement error (between test variability), **Figure 5** shows the estimated rate-of-change (least-squares slopes) at each VF location. We assume that for the 6-month study period the true rate of change was approximately zero, and so any non-zero slope estimates represent random error. This assumption is reasonable given the relatively short timeframe, that all participants were believed to be perimetrically stable, and the fact that when all four HFA tests were considered, almost as many points exhibited positive slopes (increasing sensitivity, **Fig 5A**, *red squares*) as negative slopes (decreasing sensitivity, **Fig 5A**, *blue squares*): Ratio = 0.86 (*CI*_95_ = 0.74 – 1.01; see **Fig 5C** for distribution).

When only a single (randomly selected) pair of HFA pre- and post-test results was considered (i.e., the current clinical reality following two hospital appointments), mean absolute error was 1.96 *dB* (*CI*_95_: 1.7 − 2.3; **Fig 5B**, *grey shaded region*). As progressively more home-monitoring tests were also considered (**Fig 5B**, *filled circles*) measurement error decreased to 0.35 *dB* (*CI*_95_: 0.3 − 0.4). In 37 of the 38 eyes (97%; HFA_*post*_ data missing for participant 16), mean absolute error [MAE] was smaller when home-monitoring data were included, with MAE reducing by more than 50% in 90% of eyes (Median reduction: 85%, *CI*_95_: 82 – 87%). For reference, a reduction of 20% in variability is generally considered clinically significant, and allows progression to be detected one visit earlier^49^. If we consider the home-monitoring data alone (i.e., without any HFA data included; **Fig 5B**, *unfilled squares*), measurement error was still smaller after 6 home monitoring tests (0.78 *dB*; *CI*_95_: 0.6 – 1.1) versus two HFA tests alone (1.96 *dB*), with a median reduction in MAE of 68% (*CI*_95_: 57 – 76%).

**Fig 5.**
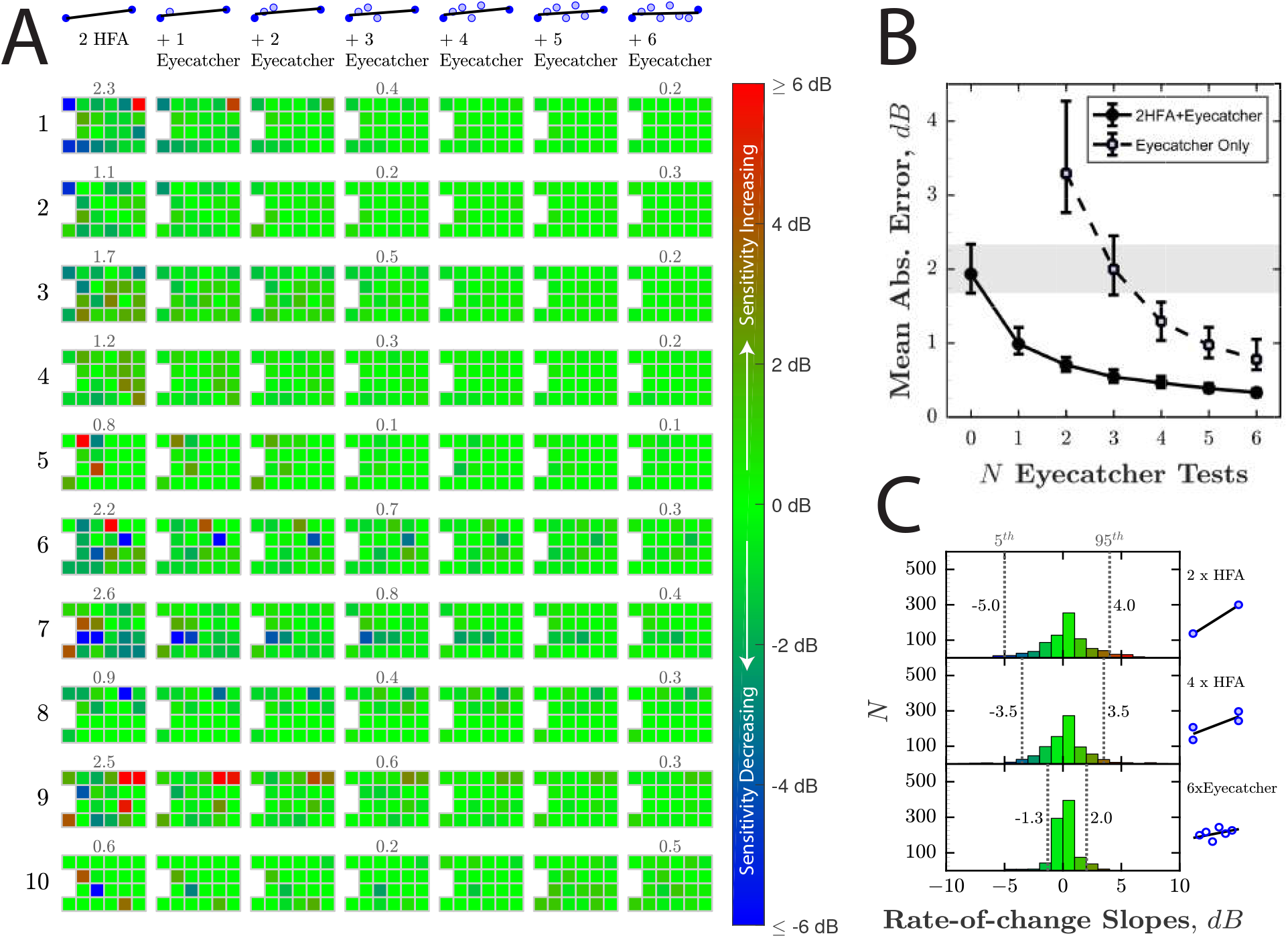
Reduction in measurement error (between-test measurement variability) from home-monitoring. **(*A*)** Estimated rate of change, as increasing numbers of Eyecatcher tests are added to a single (randomly selected) pair of HFA pre-/post-test results. As described in the main text, the true rate of change is assumed to be 0, so any non-zero values represent measurement error. Ten of 40 eyes are shown here (same eyes as **Fig 4**). Results from the remaining 30 eyes are given in **Supplemental Material S6–8**. Numbers above tests show Mean Absolute Error [MAE], which would ideally be zero. **(*B*)** Mean [*±CI*_95_] MAE, averaged across all 40 eyes, as a function of *N* home monitoring assessments (months). Filled circles correspond to the scenario in *(A)*, and show how measurement error decreased as Eyecatcher data were added to a random pair of HFA_*pre*_/HFA_*post*_ assessments (i.e., ‘Ancillary Home Monitoring Scenario’). Unfilled markers show measurement error if Eyecatcher data were considered in isolation, without any HFA data (i.e., ‘Exclusive Home Monitoring Scenario’). Error bars denote 95% confidence intervals, derived using Bootstrapping (Bias-corrected accelerated method, *N* = 20000). The shaded region highlights the 95% confidence interval (*CI*_95_: 1.7−2.3 *dB*) given only a single random pair of HFA assessments(i.e., the current clinical reality after two appointments). **(*C*)** Histograms showing the distributions of all 880 rate-of-change slopes (22 VF locations × 2 eyes × 20 participants). Vertical dashed lines show the 5^*th*^ and 95^*th*^ percentiles.

Either with or without HFA data included, there was no significant difference in MAE between the eyes of participants who received initial practice with Eyecatcher, and those who did not [*Independent samples t-test*; *P_with_* = 0.864, *P_without_* = 0.812].

In some individuals (e.g., ID3, ID13), MDs measured at home were systematically higher, in both eyes, than those measured in clinic. This difference was not significant across the group as a whole [*Repeated measures t-test of MD*: *t*_39_ = −1.08, *P* = 0.286], and may indicate individual differences in fixation stability or viewing distance. They are not likely due to ambient illumination levels, which tended to be highly variable (both within- and between-individuals), but with little apparent effect on the data (see **Supplemental Figures S9–10**).

Median test duration for Eyecatcher was 4.5 mins (*Quartiles*: 3.9 – 5.2 mins) and did not vary systematically across the 6 sessions [*F*_(5,227)_ = 0.808, *P* = 0.547; see **Supplemental Figure S11**]. For comparison, median test duration for the HFA (SITA Fast) was 3.9 *mins* (*Quartiles*: 3.3 − 4.6 *mins*), and was faster than Eyecatcher in 30 of 40 eyes (despite the HFA testing over twice as many VF locations).

## 4. Discussion

Home-monitoring has the potential to deliver earlier and more reliable detection of disease progression, as well as service benefits via a reduction in in-person appointments. Here we demonstrate, in a preliminary sample of 20 volunteers, that glaucoma patients are willing and able to comply with a monthly VF home-testing regimen, and that the VF data produced were of good quality.

98% of tests were completed successfully (adherence), and the data from 6 home-monitoring tests were in good agreement with 4 SAP tests conducted in clinic (accuracy). This is consistent with previous observations that experienced patients can perform VF testing with minimal oversight^50^, as well as with recent findings from the Age-Related Eye Disease Study 2 (AREDS2)-HOME study group, showing that home-monitoring of hyperacuity is able to improve the detection of neovascular age-related macular degeneration (AMD)^51^.

The use of home-monitoring data was shown to reduce measurement error (between-test measurement variability). When home-monitoring data were added to 2 SAP assessments made 6 months apart (the current clinical reality), measurement error decreased by over 50% in 90% of eyes. Given that a 20% reduction in measurement variability is generally considered clinically significant (i.e., allows progression to be detected one hospital-visit earlier^49^), this suggests that, even with present technology, home monitoring could be beneficial for routine clinical practice (e.g., support more rapid interventions). Furthermore, while we assume that ancillary home-monitoring, designed to supplement and augment existing SAP, would be the generally preferred model, it was encouraging that robust VF estimates were obtained even when home-monitoring data were considered in isolation. This suggests that home-monitoring may be viable in situations where hospital assessments are impractical, such as in domiciliary care, or in the wake of pandemics such as COVID-19^52^.

Home monitoring could also assist with clinical trials. For example, the recent UKGTS trial^53^required 516 individuals to attend 16 visual field assessments over 24 months: a substantial undertaking, of the sort that can make new treatments prohibitively costly to assess^54,55^. By allowing more frequent measurements of geographically diverse individuals, home-monitoring could lead to cheaper, more representative trials, and could potentially reduce trial durations (i.e., evidence treatment effects sooner).

There were, however, instances where the home-monitoring test performed poorly. In 21 tests (9%), MD deviated by more than ±3 *dB* from the median (7 deviating by more than ±6 *dB*). As has been shown previously by simulation^14^, the effects of these anomalous tests were compensated for by the increased volume of ‘good’ data. However, poor quality data should ideally be averted at-source, and it was encouraging that many of these 21 anomalous tests could be identified by applying machine-learning techniques to recordings of participants made using the tablets’ front-facing camera (see **Supplemental Figure S5**). It is also notable that when interviewed at the end of the study, some participants already suspected some tests of being anomalous (e.g., due to a long test duration, or a feeling that they had not performed well). Consideration may therefore need to be given in future as to whether participants should have the ability to repeat tests or provide confidence ratings.

Regarding adherence, one participant (ID20) was advised by the study team to discontinue home-monitoring after 4 months, after reporting that the test was compounding chronic symptoms of dizziness (though interestingly their data appeared relatively accurate and consistent up to this point; see **Fig 2**). This adverse effect was not unique to Eyecatcher, and the patient reported similar reactions following conventional SAP. However, this highlights that it may be helpful to tailor the use and frequency of home-monitoring to the needs and abilities of individual patients, in contrast to the current “one size fits all” approach to VF monitoring^12,56,57^.

### 4.1. Study Limitations & Future work

The present study is only an initial feasibility assessment, examining a small number of self-selecting volunteers. It remains to be seen how well home-monitoring scales up to routine clinical practice or clinical trials. It will be particularly important to establish that home-monitoring is sustainable over a longer periods, and is capable of detecting rapid progression^11–15^.

Cost-effectiveness of glaucoma home-monitoring has also yet to be demonstrated, and it would be helpful to perform an economic evaluation of utility, similar to that reported recently for AMD home-monitoring^58^. For this, it would be instructive to consider not just home-monitoring of VFs alone, but also in conjunction with home-tonometry, which also appears increasingly practicable^59^. Long-term, there are even signs that optical coherence tomography^60^ (OCT) and smartphone-based fundus-imaging^61–63^ are becoming easy enough to be administered by lay persons, and these might also be explored in future home-monitoring trials.

Further, it may be that focused home-monitoring — targeted at high-risk/benefit glaucoma patients — is cost-effective, even if the indiscriminate home-monitoring all patients is not^14^. Thus, it may be best to concentrate home-monitoring resources on those patients whose age^11^ or condition^17–19^ makes them most likely to experience debilitating vision loss within their lifetime. It may also be worth considering the potential secondary benefits of home-monitoring, such as improved patient satisfaction and retention^56,64^, or better treatment adherence. Thus, it is well established many glaucoma patients find hospital visits stressful and inconvenient^56,65^, and home-monitoring might be welcomed as a way of saving time, travel, and money. Treatment adherence is known to increase markedly prior to a hospital appointment^66^ (“white-coat adherence”), or when patients receive automated reminders^67,68^, and it is conceivable that the anticipation of regular home monitoring could provide a similar impetus. Following COVID-19, home-monitoring of VFs may also be desirable from a public health perspective, as a way of reducing the time each patient spends in clinic, and as a way of reducing the risk (real or perceived) of infection from conventional SAP apparatus.

### 4.2. Test Limitations & Future work

The test itself (Eyecatcher) was intended only as a proof of concept, and was crude in many respects. In fact, we consider it highly encouraging, and somewhat remarkable, that the results were as promising as they were, given the low level of technical sophistication. There are several ways in which the test could be improved in future.

In terms of software, the test algorithm (a rudimentary implementation of ZEST^41^) could be made faster and more robust: most straightforwardly by using prior information from previous tests, and by using a more efficient stimulus-selection rule^69^. The source code for the present test is freely available online for anyone wishing to view or modify it. Interestingly, while Anderson and colleagues^14^ anticipated home tests would be brief, the relatively long durations in the present study (Median: 4.5 mins per eye) were not cited as a concern by participants (although two observed that test durations were longer and more variable than conventional SAP). It may be that when it comes to home-monitoring, less focus should be placed on test duration than in conventional perimetry(i.e., given the time saved by not having to travel to and wait in clinic). Instead, focus should be directed towards usability (e.g., the ability to pause, resume, or restart tests). A full qualitative analysis of patients’ views is in preparation, and will be reported elsewhere.

In terms of hardware, the future use of head-mounted displays (or ‘smart-glasses’) would allow for widefield testing, and would obviate many practical concerns regarding uncontrolled viewing distance, improper patching, or glare from ambient lighting. These potential confounds did not appear to be limiting factors in the present study, but could be problematic in less compliant individuals, or those disposed to cheat or malinger. The future integration of eye-tracking could likewise be helpful for monitoring fixation, or for supporting eye-movement perimetry^33,70^; while iris-scanning and facial-recognition could be used to ensure that the correct eye/person is always tested. Long-term, test data will need to be integrated securely into medical records systems, and consideration given how to maintain accurate screen calibrations over extended periods of use^44^.

## Data Availability

Data available upon request.

## Supplemental Material

### 1. Additional *Methods*

**Fig S1.**
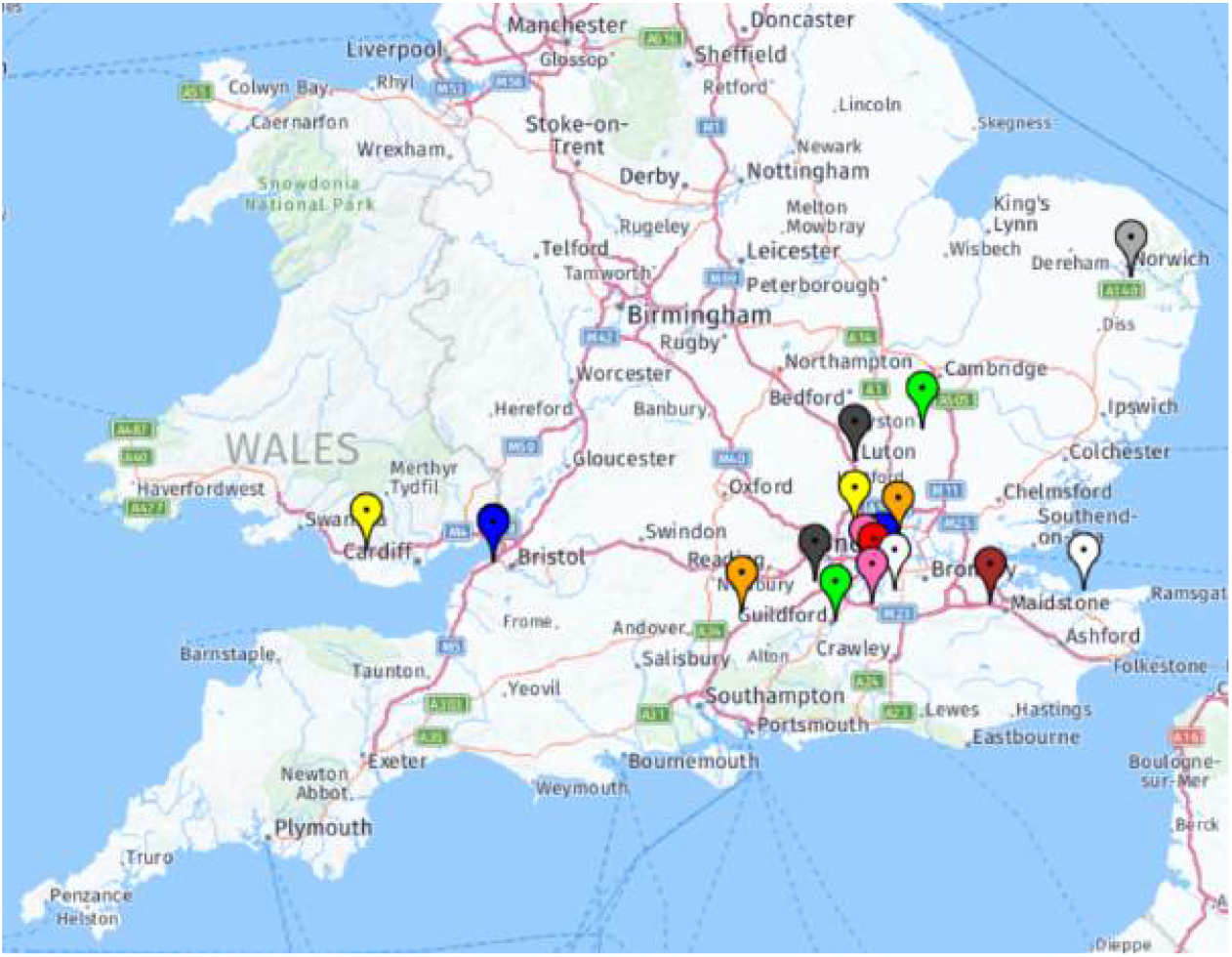
Geospatial distribution of participants.

### 2. Additional *Results*

**Fig S2.**
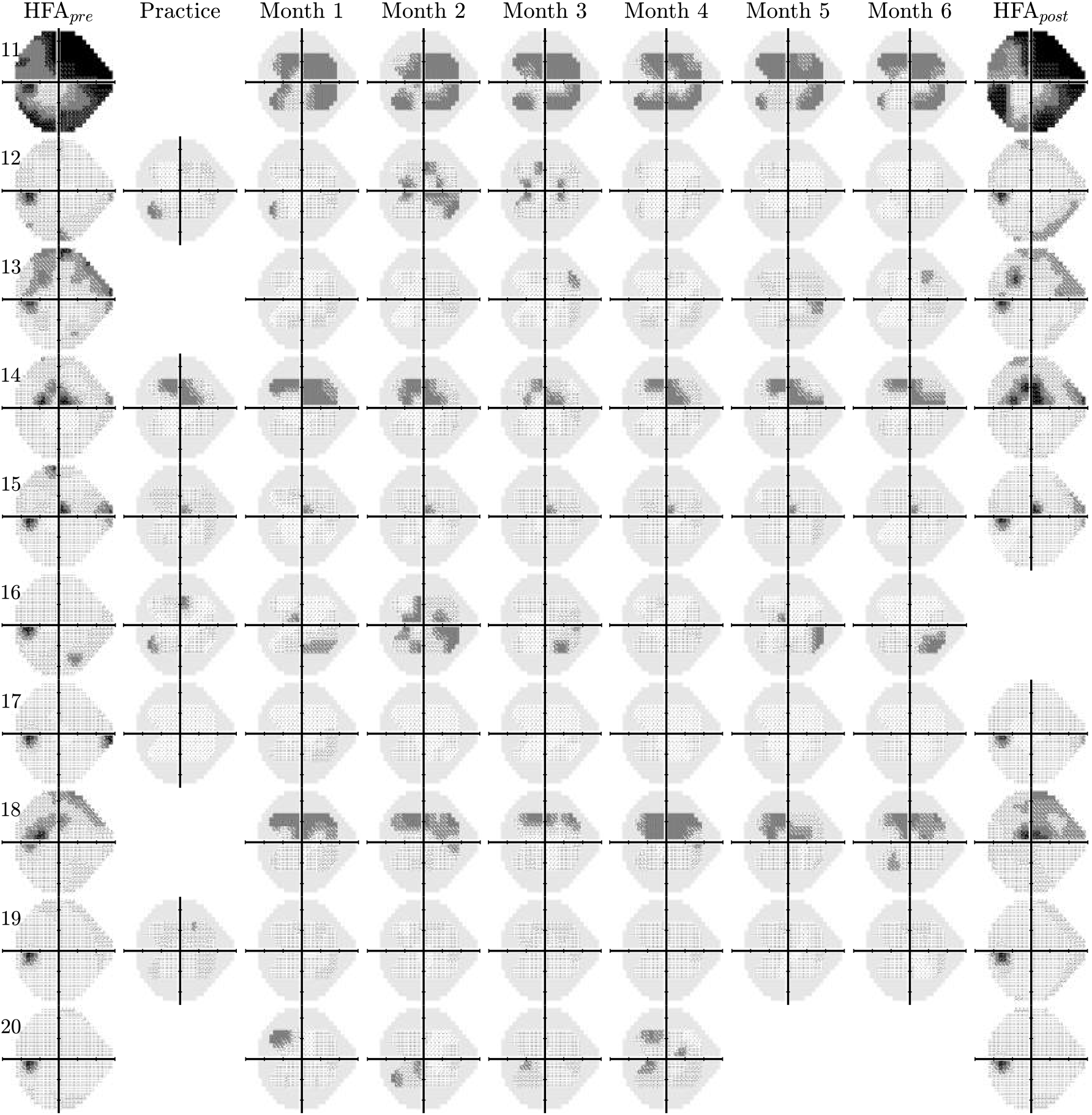
Raw right eye VF data for participants 1–10 (same format as left-eye data, shown in Figure 4 of *Main Manuscript*).

**Fig S3.**
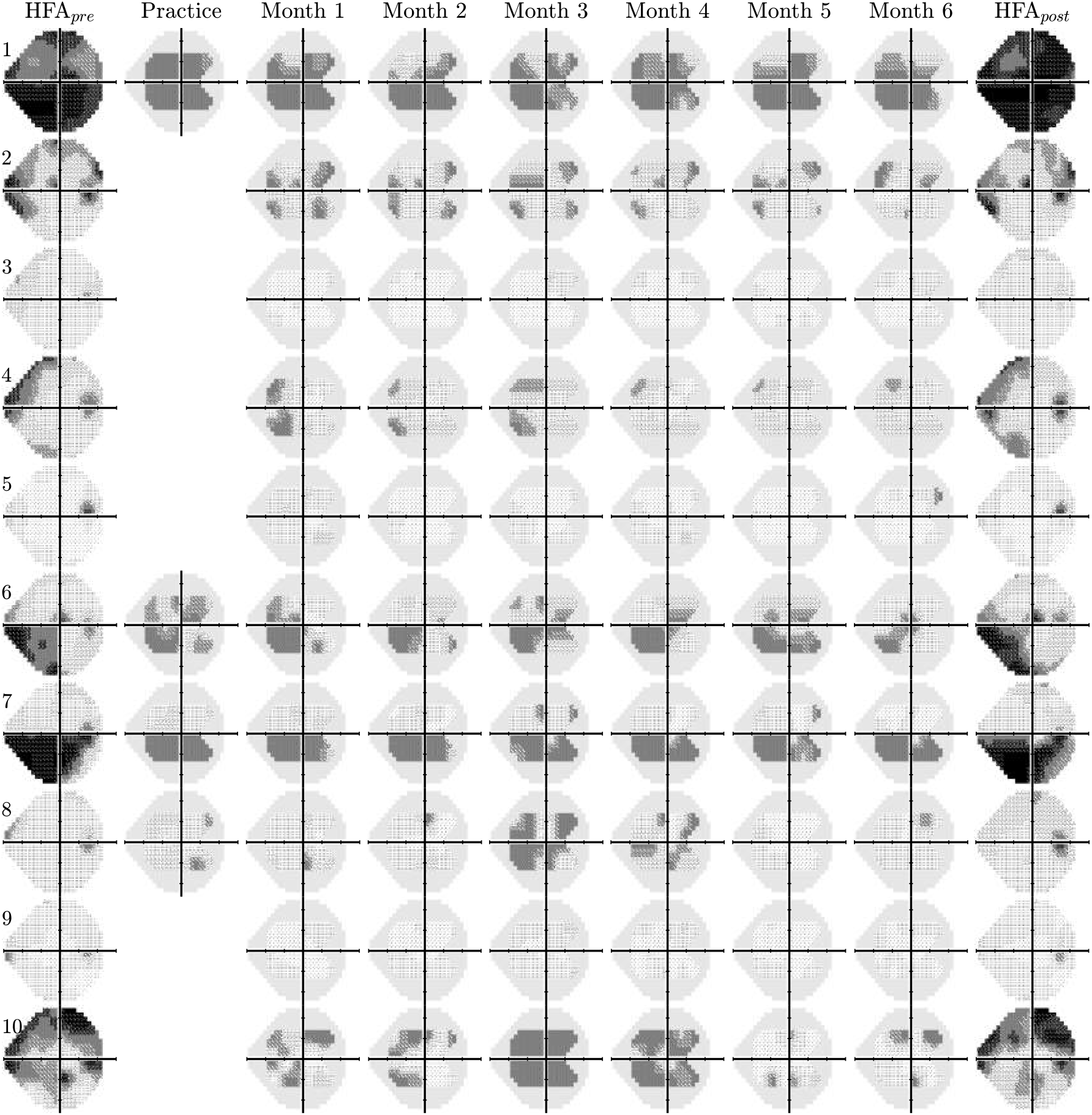
Raw left eye VF data for participants 11–20 (same format as Figure 4 of *Main Manuscript*).

**Fig S4.**
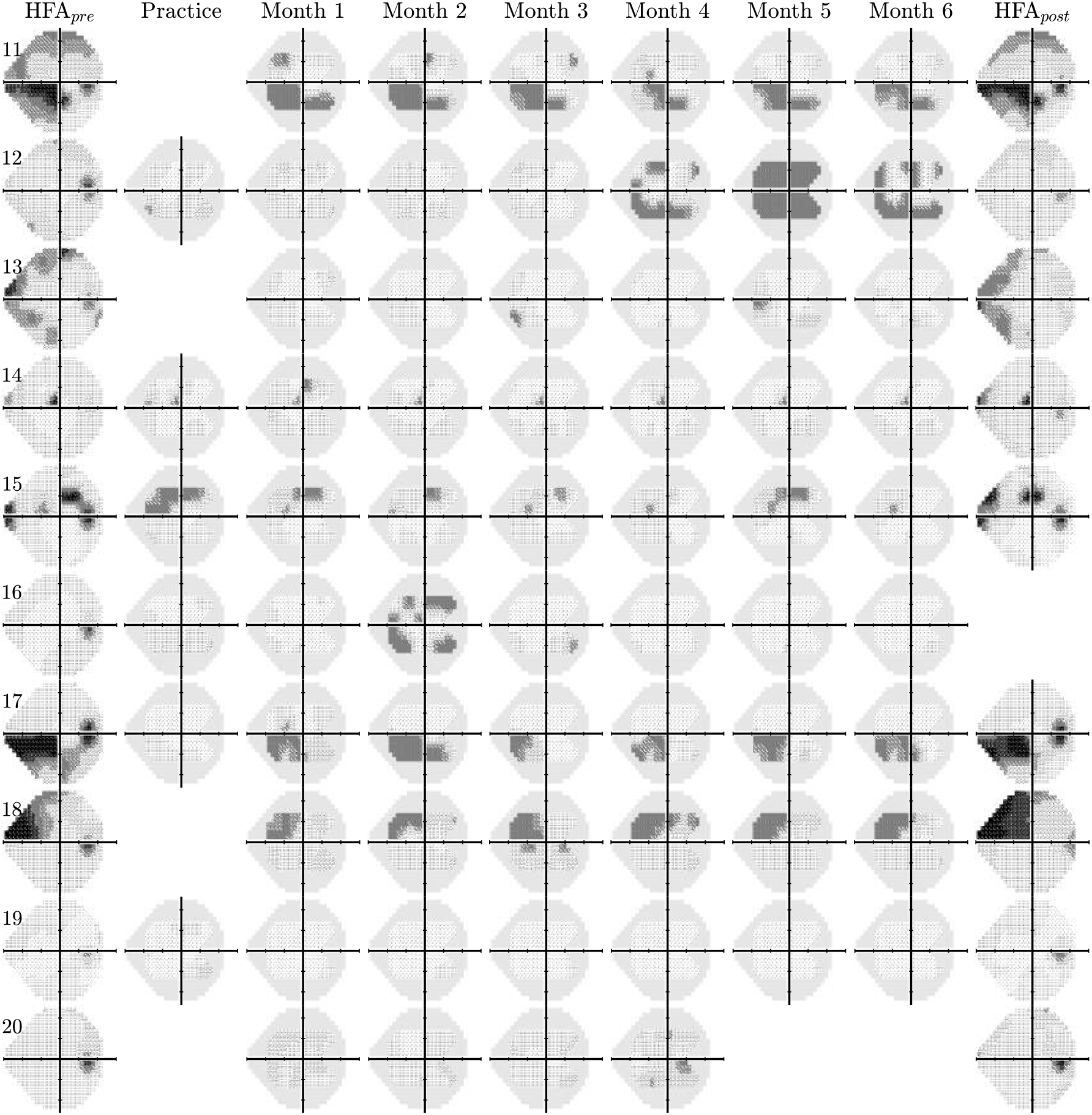
Raw right eye VF data for participants 11–20 (same format as Figure 4 of *Main Manuscript*).

**Fig S5.**
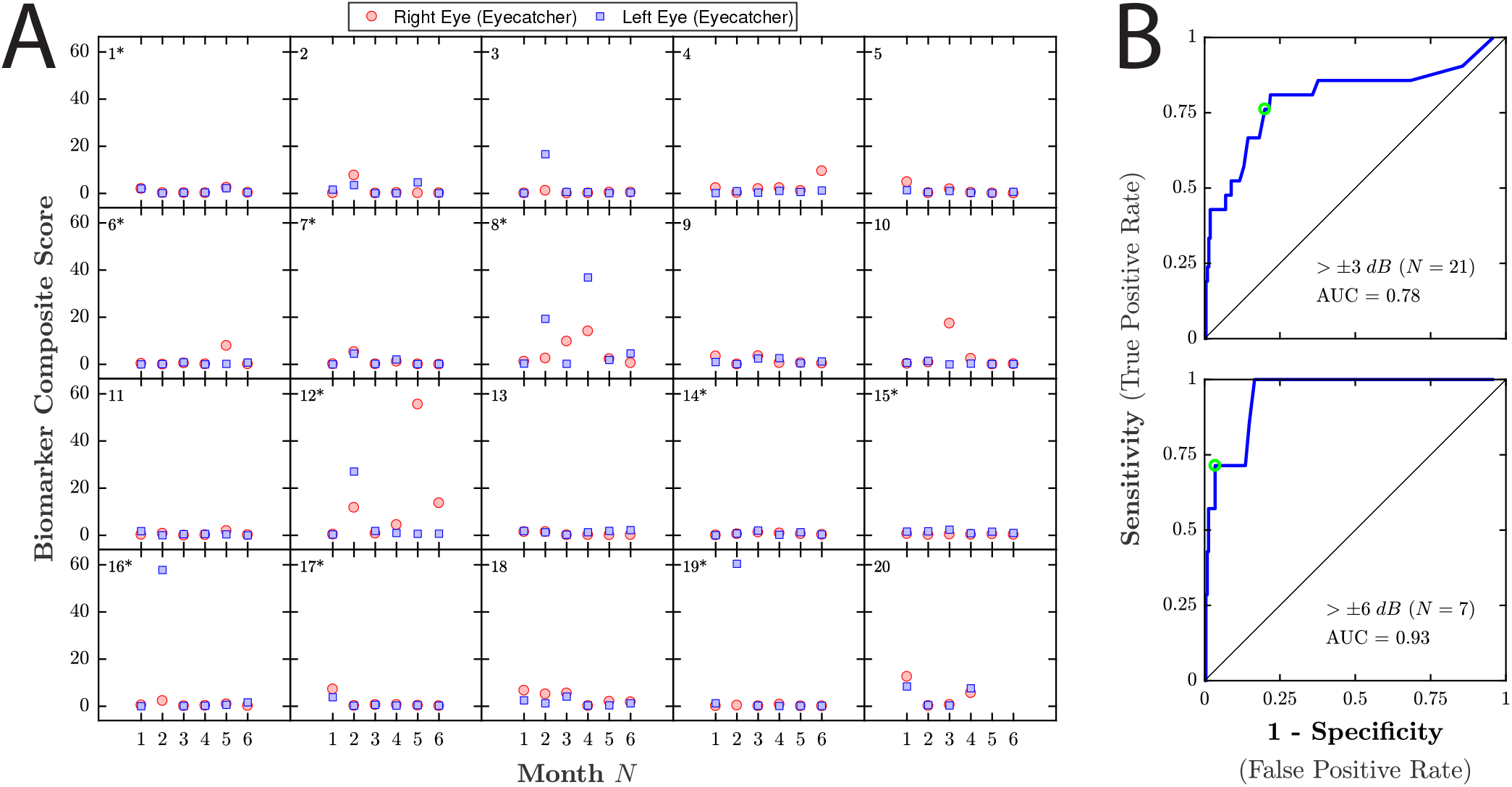
Data from the tablet’s front-facing camera (and participant response latencies) were used identify anomalous tests. Following a method similar to *Jones et al, TVST, 2020*, a “Biomarker Composite Score” (BCS) was computed by linearly combining three variables: (1) mean response latency; (2) Variability in camera pixel intensity throughout the test (i.e., which would be high if somebody suddenly turned on a light or opened a door); (3) an OpenFace facial expression metric. Formally, 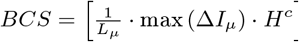, where *L_μ_* is mean response latency, in seconds, across all trials, Δ*I_μ_* is the difference in mean pixel intensity between consecutive video frames, *H* is estimated Happiness (a value from 0–1, from OpenFace), and *c* is the estimated confidence of H (a value from 0–1, from Open-Face). ***(A)*** Raw BCS for each individual test, for comparison with the MD scores shown in the same format in Figure 2 (*Main Manuscript*). ***(B)*** Corresponding Receiver Operating Characteristics (ROCs). Moderately anomalous test results (MD more than ±3 *dB* from the median) could be identified with reasonable sensitivity/specificity (*AUC* = 0.78). Highly anomalous test results (MD more than ±6 *dB* from the median) could be identified with high sensitivity/specificity (*AUC* = 0.94). In principle these tests could be excluded or down-weighted post-hoc, or individuals could be automatically invited to repeat such tests.

**Fig S6.**
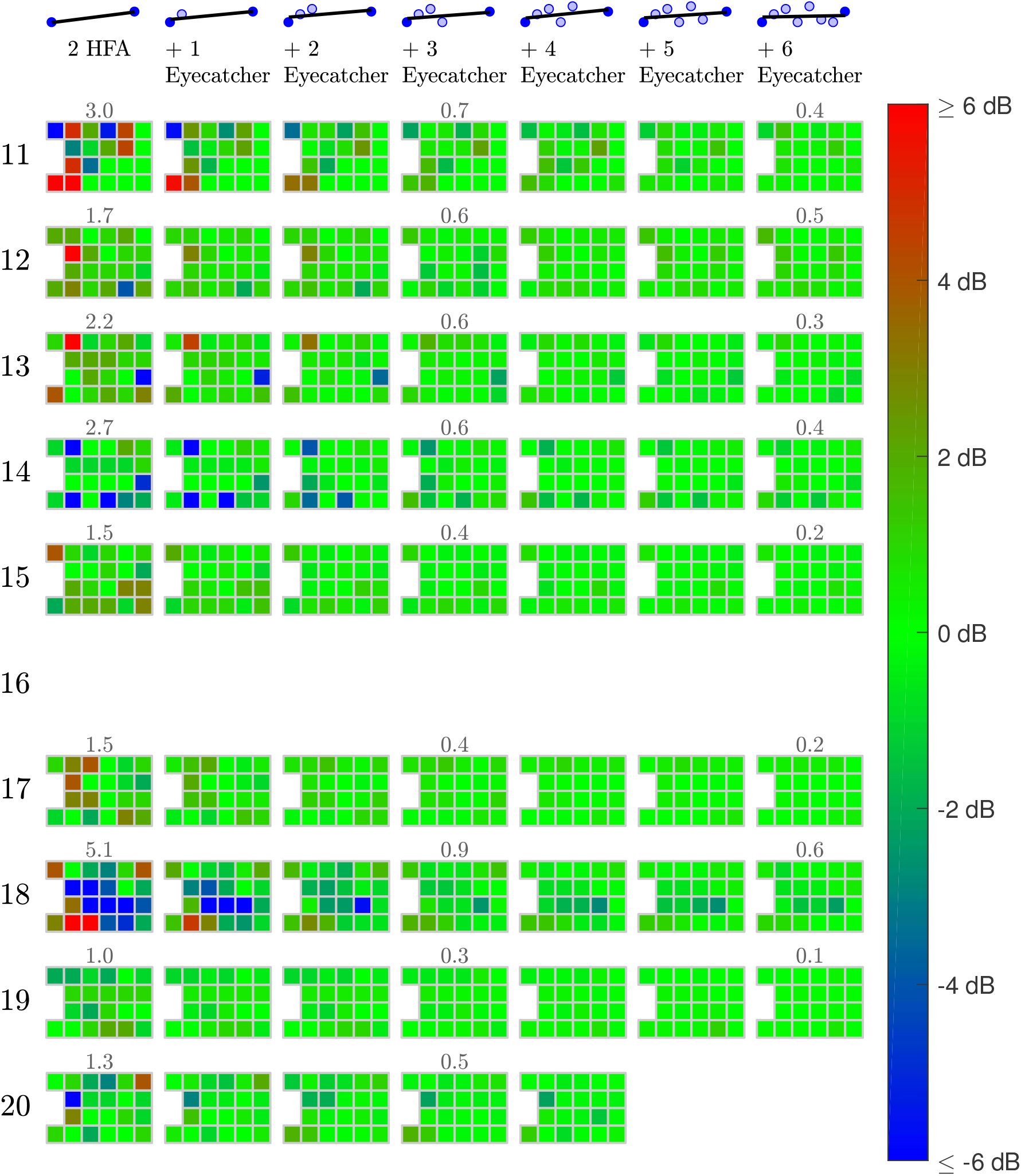
Benefit of home monitoring (reduction in rate-of-change measurement error) for the right eyes of participants 1–10 (same format as Figure 5A of *Main Manuscript*).

**Fig S7.**
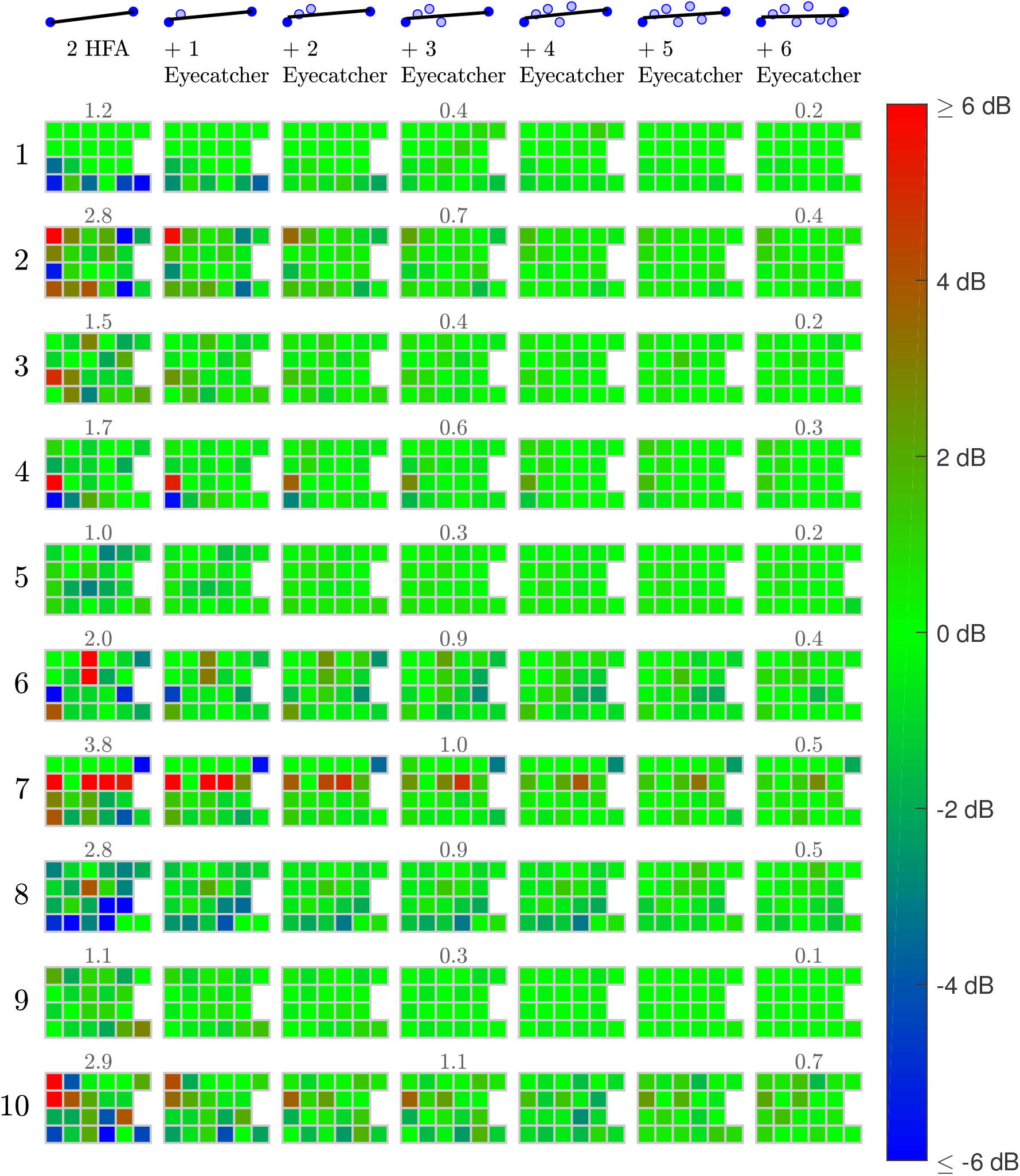
Benefit of home monitoring (reduction in rate-of-change measurement error) for the left eyes of participants 1–10 (same format as Figure 5A of *Main Manuscript*).

**Fig S8.**
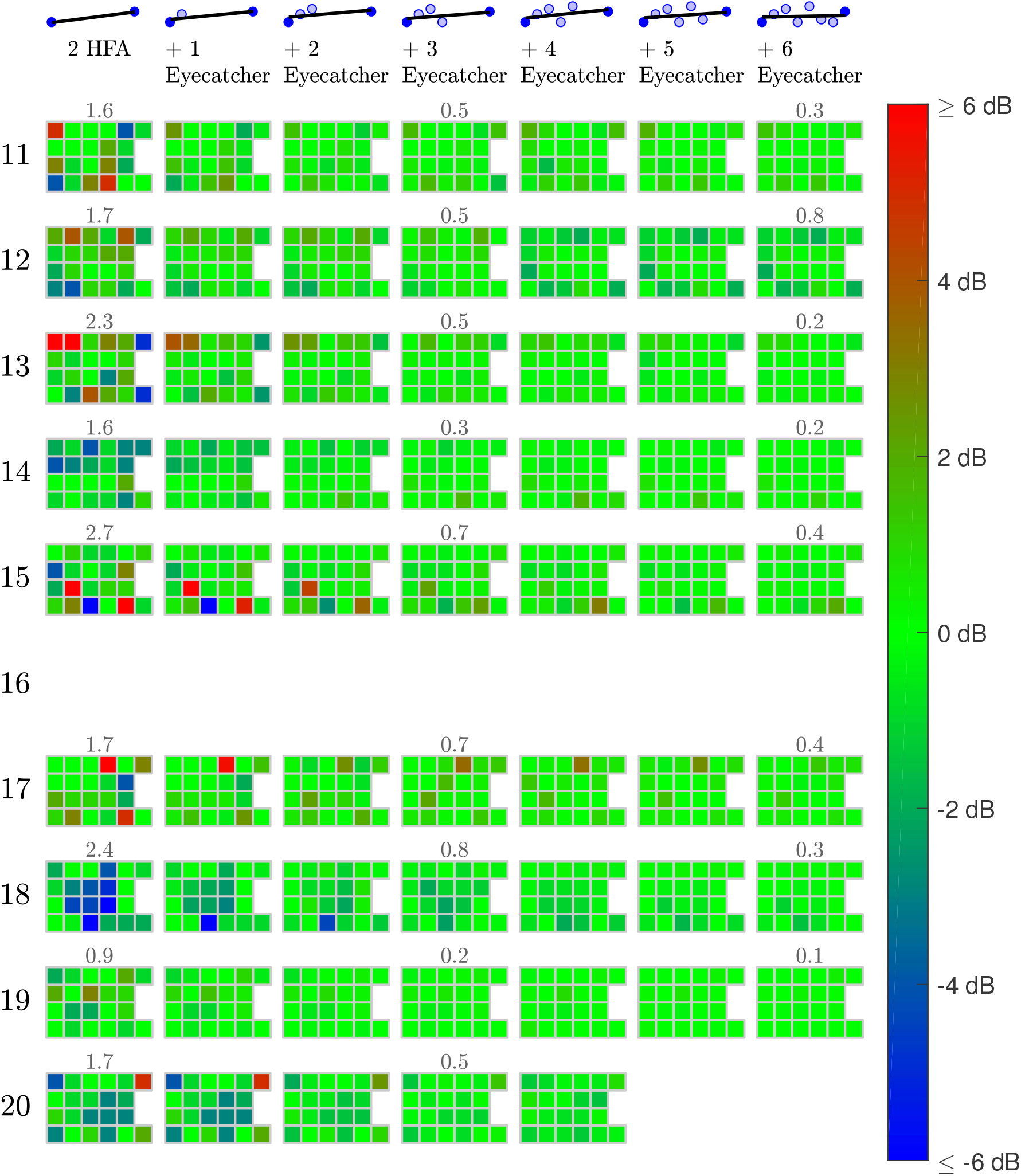
Benefit of home monitoring (reduction in rate-of-change measurement error) for the right eyes of participants 1–10 (same format as Figure 5A of *Main Manuscript*).

**Fig S9.**
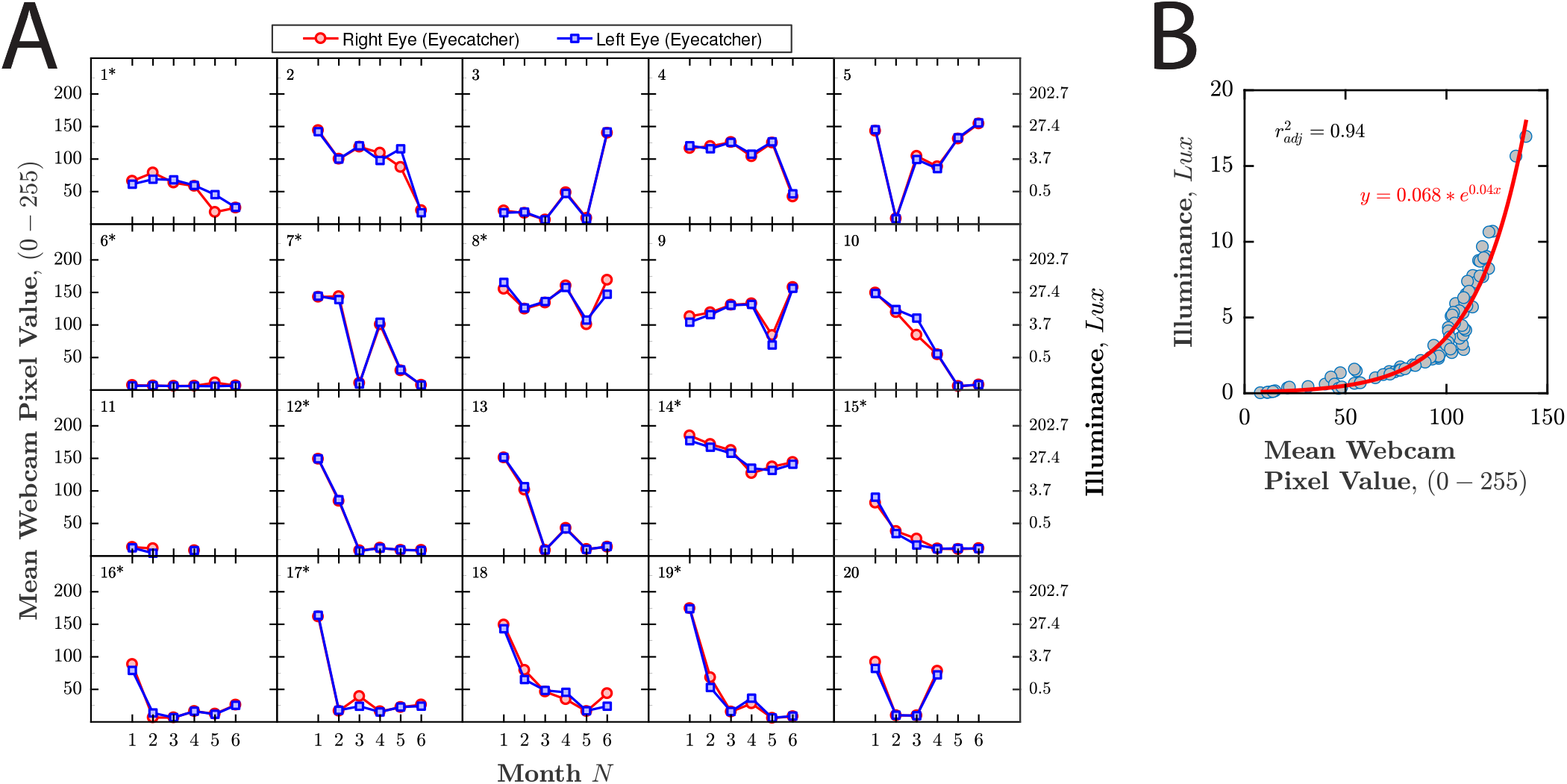
Ambient illumination levels during home testing. ***(A)*** Each marker represents the mean webcam pixel intensity value, averaged across all pixels and all frames. A secondary *y*-axis shows the corresponding estimated illuminance, in *Lux*, based on a the calibration data shown in (B). Note that for participant 20, data are missing for tests 5 and 6 as they stopped performing the test. For participant 11, data are missing for tests 3, 5, 6 because they chose to cover the camera on these tests. When questioned subsequently, they stated that this was because they found the recording light distracting (not for privacy). ***(B)*** Calibration data. Each marker (*N* = 90) represents raw measurements, made using an AOPUTTRIVER AP-881D Digital Lux Meter. The red line is the best fitting (least-square) exponential function, which provides an acceptable approximation of the data for present purposes.

**Fig S10.**
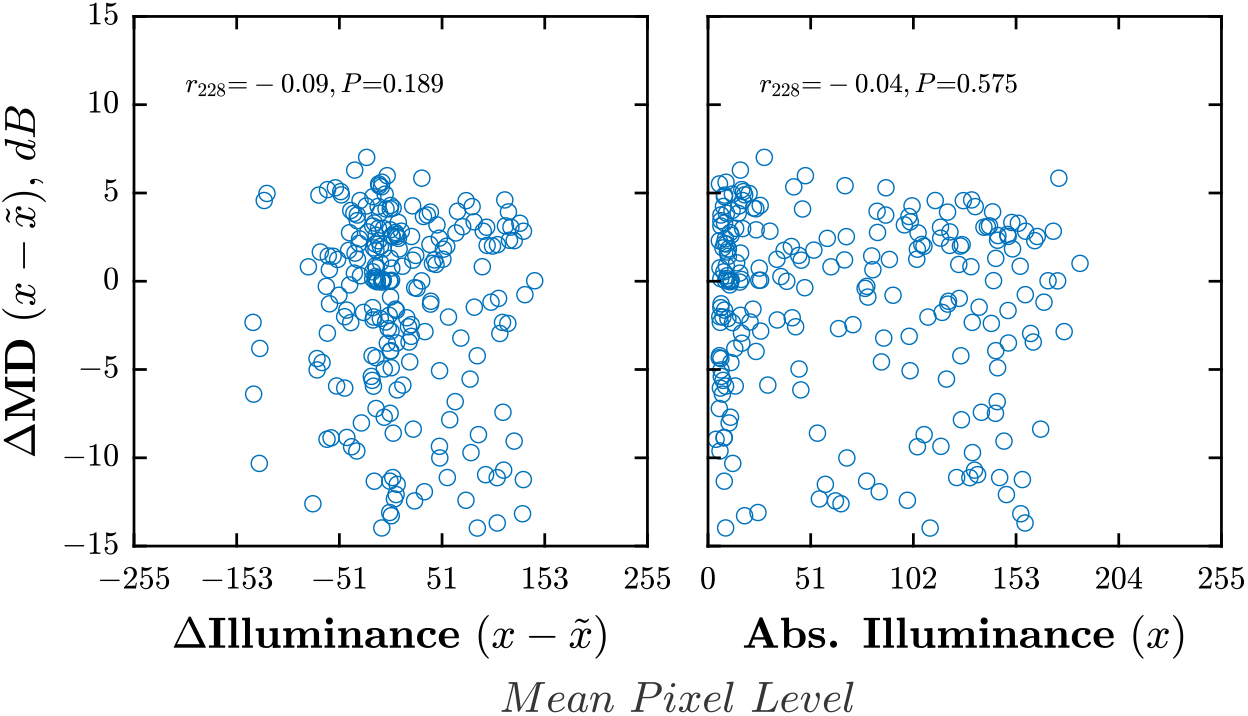
Relationship between ambient lighting and estimated VF loss. ***(A)*** There was no relationship between changes in illuminance and changes in test score [*r*_214_ = 0.07, *P* = 0.320]. ***(B)*** There was no relationship between absolute illuminance and absolute test score [*r*_214_ = −0.06*, P* = 0.373].

**Fig S11.**
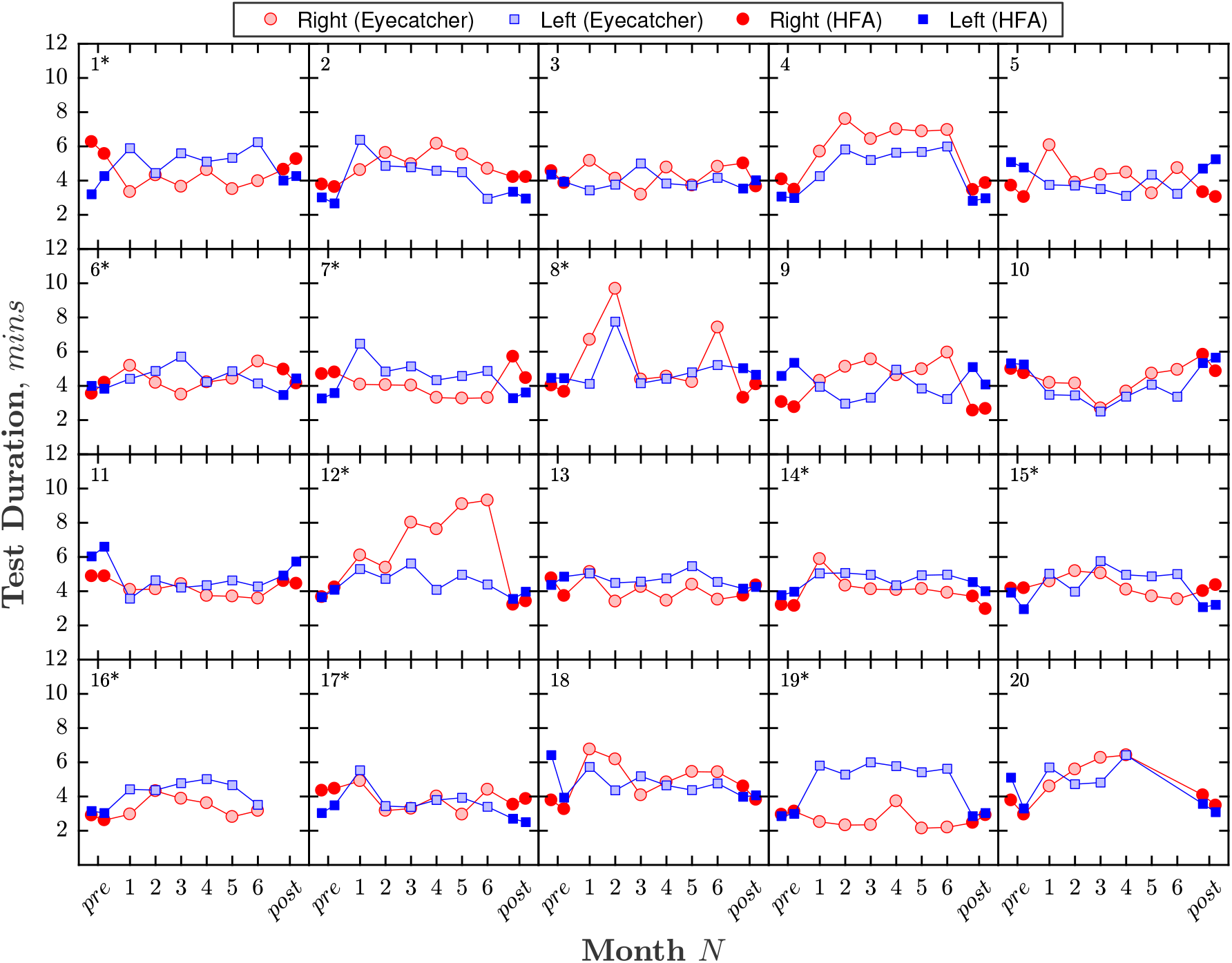
Test duration. Same format as Figure 2 of *Main Manuscript*.

**Fig S12.**
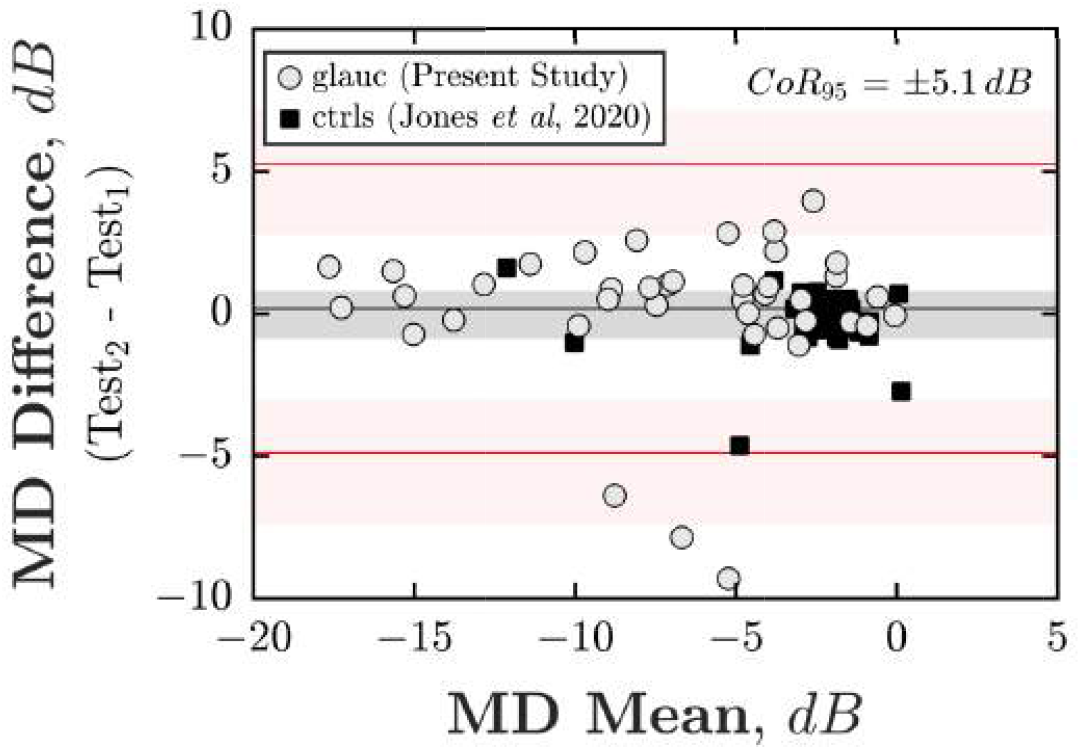
Reliability of each individual home-test (test-retest repeatability). Bland-Altman agreement from the first two Eyecatcher home tests (Month 2 – Month 1). Each marker corresponds to a single eye. Solid squares show analogous data from 46 control eyes, reported previously by *Jones et al, TVST, 2020*. These data are shown for illustration only, and were not included when computing the limits of agreement shown. If included, the 95% Coefficient of Repeatability was ±2.8 *dB*.

## Notes

### Competing Interest Statement

The authors have declared no competing interest.

### Funding Statement

This study was funded by a Fight for Sight (UK) project grant (#1854/1855) and by the International Glaucoma Association/College of Optometrists 2019 Award (which is funded by the IGA and administered by the IGA in conjunction with the College of Optometrists). Author DA was supported by the European Union's Horizon 2020 research and innovation program under Marie Sklodowska-Curie grant agreement No. 675033. The funding organizations had no role in the design or conduct of this research.

### Author Declarations

The study was approved by the Ethics Committee for the School of Health Sciences, City, University of London (#ETH1819-0532), and carried out in accordance with the tenets of the Declaration of Helsinki.

